# Practitioner Psychologists’ Experiences of Delivering a Cognitive Behavioural Therapy Informed Crisis Intervention for Psychosis in Inpatient Settings: A Mixed Methods Investigation within the CRISIS study

**DOI:** 10.1101/2025.10.27.25338870

**Authors:** Amy Robinson, Nicola Morant, Aderayo Ariyo, Hannah Butterworth, Patrick Nyikavaranda, Nira Malde Shah, Ceri Dare, Karen Persaud, Ella Guerin, Mary Birken, Sonia Johnson, Lisa Wood

## Abstract

**Objectives:** Cognitive behaviour therapy for psychosis (CBTp) should be delivered in psychiatric inpatient settings, yet little is known about therapists’ perspectives on delivering it. This study examined therapists’ perspectives on delivering a crisis-focused CBTp-informed (cCBTp) intervention.

**Design:** The study was part of the CRISIS (CRISis cbtp in Inpatient Settings) study, a feasibility randomised controlled trial of the cCBTp intervention for inpatients experiencing psychosis. A mixed methods approach combined therapy log data and qualitative interviews with therapists after the trial therapy was complete.

**Methods:** Seven CRISIS study therapists completed a therapy log, which we analysed descriptively. Semi-structured interviews with six of these therapists explored their experiences with intervention training and delivery, which was analysed using thematic analysis.

**Results:** The results from the therapy log demonstrated that therapists’ undertook a comprehensive assessment and prioritised engagement with all participants, and most developed a formulation, which informed change strategy delivery aligned with the patient’s goals. In the qualitative interviews, therapists emphasised the importance of delivering culturally competent flexible, person-centred therapy and supporting patients to work towards goals such as coping with the crisis and discharge planning. They described challenges of delivering therapy in the acute crisis context including interruptions to therapy sessions, patients experiencing acute symptoms, and environment restrictions.

**Conclusions:** The study demonstrated the importance of delivering cCBTp collaboratively and supporting patients in understanding and managing their own crisis. It also identified several challenges therapists had delivering the therapy. Further research is needed to explore therapists’ experiences of delivering psychological interventions in this setting.

**Practitioner Points:**

1. Therapists must be flexible in their approach and remain adaptable with session timing, location and content to accommodate disruptions in inpatient settings, ensuring continuity of therapy despite high levels of distress and non-attendance.
2. A validating, therapeutic relationship is central to support patients with psychosis in crisis, enabling trust, safety and effective goal-directed work.
3. Therapists and patients collaboratively developing a crisis-focused formulation helps patients make sense of their current crisis, facilitates empowerment and enhances relapse prevention and discharge planning.
4. Therapist should actively explore and integrate patient’s cultural experiences including experiences of racism into therapy to help strengthen engagement and provide a holistic understanding of a patients’ experiences.

## Introduction

Acute mental health inpatient settings provide intensive care to patients experiencing severe mental health crises, often characterised by distressing symptoms and heightened risk of harm to self, to others, and/or from others (NHS England, 2023). The Long-Term Plan (NHS England, 2019) emphasises the need for therapeutic inpatient environments, while the Mental Health Task Force (NHS England, 2016) advocates for timely, evidence-based psychological therapies in these settings. Inpatient care should be holistic, based on a biopsychosocial framework (Bowers, 2014), and delivered by a multidisciplinary team (MDT) that includes psychologists providing therapy (Royal College of Psychiatrists, 2015). The National Institute of Health and Care Excellence guidelines (NICE, 2014) recommend offering psychological therapy to all inpatients and many express a strong preference for it (Berry et al., 2022). Nonetheless, research shows that reliance on medication and involuntary admission remain common in inpatient settings, with limited access to psychological interventions (Radcliffe & Bird, 2016; Voskes et al., 2021). Additionally, patients from ethnic minority backgrounds, particularly Black African and Black Caribbean groups are overrepresented in inpatient settings and are nearly five times more likely to be detained under the Mental Health Act (NHS Digital, 2024). This disparity is further underscored by the weighted prevalence of psychosis, which is 14.2 per 1,000 in Black ethnic minority populations compared to 4.4 per 1,000 in the white population (Qassem et al., 2015). Despite this increase in prevalence and associated risk, these groups often have less access to psychological therapies (Schlief et al., 2023) and worse experiences with mental health care (Halvorsrud et al., 2018).

Cognitive Behaviour Therapy for psychosis (CBTp) is a psychological therapy that aims to target the distress associated with psychotic experiences by adapting the way an individual thinks and behaves. The NICE guidelines recommend offering CBTp in inpatient settings during the acute period of psychosis (NICE, 2014), which is particularly relevant given that two-thirds of individuals admitted to inpatient care are experiencing psychosis (NHS Benchmarking, 2016). Although some concerns have been raised about the potential overstatement of CBTp’s effectiveness (McKenna & Kingdon, 2014), evidence supports its efficacy in inpatient settings (Wood et al., 2020). However, uncertainty remains regarding how therapists deliver CBTp in inpatient settings. Providing therapy in these environments is complex, as patients are often involuntarily detained, experience distressing symptoms, and are on high doses of medication (Evlat et al., 2021). Furthermore, current guidelines for CBTp typically recommend 16 to 24 weekly sessions (NICE, 2014) which is impractical in inpatient settings where the average length of stay in 2023 was 39 days (Kings Fund, 2024). This highlights the critical need for a thorough evaluation of psychological interventions such as CBTp tailored to inpatient settings with a focus on crisis reduction.

The evidence for the effectiveness of CBTp in inpatient settings is limited, with only three systematic reviews being conducted which demonstrated moderate-to-poor quality. (Jacobsen et al., 2018; Paterson et al., 2018; Wood et al., 2020). Improvements were seen in anxiety, depression, and readmission rates; however, improvements in psychotic symptoms did not persist at follow-up (Paterson et al., 2018; Wood et al., 2020). Additionally, therapies were not tailored to the specific needs of inpatients or co-produced with key stakeholders (Jacobsen et al., 2018; Wood et al., 2022a). While CBTp is recommended for use in inpatient settings (NICE, 2014), most studies on its effectiveness have focused on community samples or outdated inpatient environments. In response to the lack of evidence regarding delivering CBTp in inpatient settings, a culturally competent, CBTp-informed crisis-focused intervention was developed in accordance with the Medical Research Council’s guidelines for complex interventions (Skivington et al., 2021). The intervention’s feasibility and acceptability were evaluated in a randomised controlled trial (RCT) known as CRISIS (CRIsis cbtp in Inpatient Settings) study, with detailed methods outlined in the protocol paper (Wood et al., 2022b). To ensure that the intervention meets the needs of patients effectively, the content and delivery process were co-produced with key stakeholders, including individuals with lived experience of psychosis and inpatient care (including service users from ethnic minority backgrounds), family and carers, and clinicians.

The crisis-focused CBTp-informed intervention (cCBTp) is based on a modular CBTp protocol designed to provide approximately six to eight therapy sessions for participants. The intervention consists of several modules (Wood et al., 2024a): engagement, assessment, and identifying priorities; crisis formulation; stabilisation, safety, and problem-solving; creation of crisis plans and crisis cards; change strategies focusing on crisis appraisals and discharge and relapse planning (see table 1). The first two modules are essential core modules (engagement, assessment, and identifying priorities and crisis formulation) and the remaining ones are applied flexibility depending on the patients’ needs. The intervention was delivered by practitioner psychologists with prior experience of delivering standard CBT in inpatient settings. The feasibility RCT of the cCBTp intervention found that the research procedures and intervention were feasible measured by recruitment and retention rates in the trial (Wood et al., 2025). Notably, patients who received the crisis-focused CBTp-informed intervention participated in qualitative interviews, revealing that they found the therapy acceptable and beneficial (Wood et al., 2024b). Examination of therapy data and therapists’ perspectives will add further depth to these findings.

**Table 1.** Outline of therapy modules.

| **Module** | **Description** |
| --- | --- |
| **Engagement, assessment, and identifying priorities** | The therapist should build a therapeutic relationship and collaboratively assess the patient's presenting crisis and reasons for admission. The therapist should identify crisis-focused goals that improve safety, reduce risk, and facilitate discharge. |
| **Formulation of the crisis** | A collaborative formulation should be developed of the patient's presenting crisis drawing on the model outlined in Wood et al., (2025) underpinned by Morrison (2017). |
| **Stabilisation, safety, and problem solving** | Intervention strategies that promote patient safety and stabilisation should be collaborative employed. This will include coping strategy development to manage emotional distress and physiological difficulties, e.g. sleep. |
| **Crisis plans and crisis cards** | Crisis plans and cards include a collection of strategies for the patient and their network to manage their crisis experiences in the moment. This will include identification of early warning signs, coping strategies and family/carer support. |
| **Change strategy work and focussing on crisis appraisals and safety behaviours** | Drawing on the Morrison (2017) protocol, adapted change strategies will be utilised to facilitate the goals identified in module 1. This can include normalisation, activity scheduling and behavioural experiments. |
| **Discharge, relapse planning, and recovery toolkits** | Bringing everything covered in therapy into a discharge plan and ensuring any new strategies/approaches, e.g. coping strategies and crisis plans, are adapted to the home environment. |

To evaluate the acceptability of the cCBTp intervention in inpatient settings, detailed and transparent reporting of therapy sessions is essential. Accurate documentation of therapeutic techniques and activities supports best practice guidelines and enables evaluation, replication, and improvement of the intervention (Hoffman et la., 2014).

Moreover, staff perspectives are crucial in examining the effectiveness of therapeutic interventions. To the author’s knowledge, only two qualitative studies have explored psychologists’ experiences of delivering therapies in inpatient settings (Small et al., 2018; Wood et al., 2019). These studies highlighted the importance of establishing a ‘human’ relationship as a foundation for therapeutic work in these environments. Furthermore, providing therapy in inpatient settings presents unique challenges that necessitate adaptations, such as flexible patient sessions and inclusive engagement (Wood et al., 2019). A significant aspect of these challenges includes the need for effective training that equips therapists with the skills and strategies to deliver therapies in such complex environments.

While these studies offer valuable insights into routine practice, they reveal a significant gap in research regarding therapists’ experiences with the training and delivery of protocol-driven crisis-focused CBTp-informed intervention within the context of a research trial.

The broad research question guiding this work was to examine the therapists’ experiences of the intervention delivery. Specifically, it aimed to explore what the therapists delivered and what their experiences were with the training for the intervention and the actual delivery of therapy.

## Method

### Reporting and ethics

The CRISIS trial was registered on the ISRCTN trial registry (ISRCTN59055607). Full Health Research Authority (HRA) and NHS Research Ethics Committee (REC) approval was granted (IRAS ID: 272043; 20/LO/0137/AM01) and the study is sponsored by the University College London. For the qualitative aspect of the study, methods are reported according to the Consolidated Criteria for Reporting Qualitative Studies (COREQ) guidelines (Tong et al., 2007).

### Study Design

This study utilised a mixed methods approach to examine the therapists’ experiences of the cCBTp intervention. This study was part of a single centre individually randomised, parallel-group, feasibility RCT design. In the CRISIS trial, 52 participants were recruited across eight wards and were randomly allocated to either treatment as usual (TAU) or cCBTp plus TAU in a 1:1 ratio. 26 were randomly allocated to cCBTp plus TAU and 26 to TAU. More detail about the trial can be found in Wood et al., (2025).

### Participant Sampling

A total of seven therapists delivered therapy in the CRISIS study. The inclusion criteria for these therapists for this current study were as follows, they must:

a. Be a Health Care and Professions Council (HCPC) registered practitioner psychologist
b. Have experience of delivering the cCBTp therapy as part of the trial
c. Be employed by the recruiting NHS trust at the time of therapy delivery.

Six of these therapists, who provided therapy to a total of 18 participants, were interviewed to explore their experiences. One therapist (LW) who provided therapy to eight patients was excluded to prevent potential bias, as they were the chief investigator of the study and involved in developing and delivering the training and intervention.

### Materials

A therapy-specific data log, based on the TIDieR checklist (Hoffmann et al., 2014), was created for therapists to record the content and delivery of each session. This log included detailed information on the number of sessions attended, the number of sessions declined or missed, the session agenda, and the types of cCBTp and non-specific cCBTp strategies used. Alongside this, a random 10% of all therapy sessions were rated using the Cognitive Therapy Rating Scale (CTRS; Blackburn et al., 2001) to ensure adherence to the CBT model (operationalised as a score of 3 or more on each CTRS item). All therapists had at least one session check for fidelity and to check self-reported data was accurate.

A semi-structured interview guide was developed with the research group in partnership with the coproduction team. The coproduction group, comprising individuals with lived experience of psychosis and inpatient care (including service users from ethnic minority backgrounds), family and carers, and clinicians, reviewed draft interview questions developed by LW and a qualitative expert (NM). The coproduction group discussed key areas, reviewed the wording of the questions, identified any missing elements, explored potential prompts, and considered other important aspects to include in the interview. For the full interview schedule, see the appendix.

### Procedure

All therapists were asked to complete the therapy-specific data log following each session, contact with the patient, or any type of indirect work. A research assistant emailed the six therapists, inviting them to complete a semi-structured interview once they had finished all of their therapy cases. Written informed consent was obtained from all participants before the qualitative interviews. The interviews were conducted remotely via Microsoft Teams by two research assistants (HB and AA) between 26th May 2022 and 15th June 2022. The interviews lasted an average of 36.74 minutes (range 30.10 to 44.02).

### Analysis

The therapy-specific data recorded in the log was descriptively analysed, including session attendance (total number of sessions attended, missed, or declined) and key characteristics including average therapy session length and location of the session. We also examined which of the cCBTp modules and non-specific cCBTp strategies including indirect work, family involvement, assistant psychologist involvement, and MDT involvement were used during therapy for each participant. This provided the total frequency of each module or strategy used at the participant level e.g. the number of participants that had therapy sessions focused on assessment.

The qualitative interviews were recorded via Microsoft Teams, transcribed verbatim, and analysed using thematic analysis (Braun & Clark, 2006) within NViVo software. Thematic analysis was conducted from a critical realist perspective, integrating both inductive and deductive methods. This approach aimed to explore therapists’ experiences with delivering cCBTp and to examine their perspectives on the acceptability of the intervention. Initially, each recorded interview was watched at least twice, and each transcript was read and reread to ensure that AR was fully immersed in the data. Initial codes were generated by coding the data line by line. These codes were then collated across interviews and grouped to form topic domains and sub-themes. Topic domains were explored across the dataset, focusing on both commonalities and variations in therapists’ perspectives.

To enhance the validity of the findings, regular collaborative meetings were held with AR’s supervisor to discuss emerging codes, themes, and patterns across the data. These discussions critically examined similarities and differences across the data, helping to further conceptualise and develop the themes.

### Research Team & Reflexivity

The research team and co-production team consisted of people from diverse backgrounds, encompassing a wide range of ages, ethnicities, genders, cultures, and professions. LW, who led on the interview schedule development, is a clinical psychologist and was principal investigator and therapist on the CRISIS study and has over 15 years of experience of working clinically in inpatient settings. AR who led on the thematic analysis and wrote this manuscript, is a research assistant in the NHS with over five years’ experience in mental health settings. During initial coding and theme development, AR maintained a reflexive journal to document biases, assumptions, and analytical decisions, thereby enhancing transparency and fostering critical self-awareness that grounded interpretations and prompted ongoing reflection on personal influence.

## Results

### Therapist Demographics

Table 2 presents the demographics of the therapists whose interviews were analysed for the thematic analysis. All participants were female, the majority were clinical psychologists, and most had at least three years of experience working in inpatient settings, with all having a minimum of three years of experience working with individuals with psychosis.

**Table 2.**
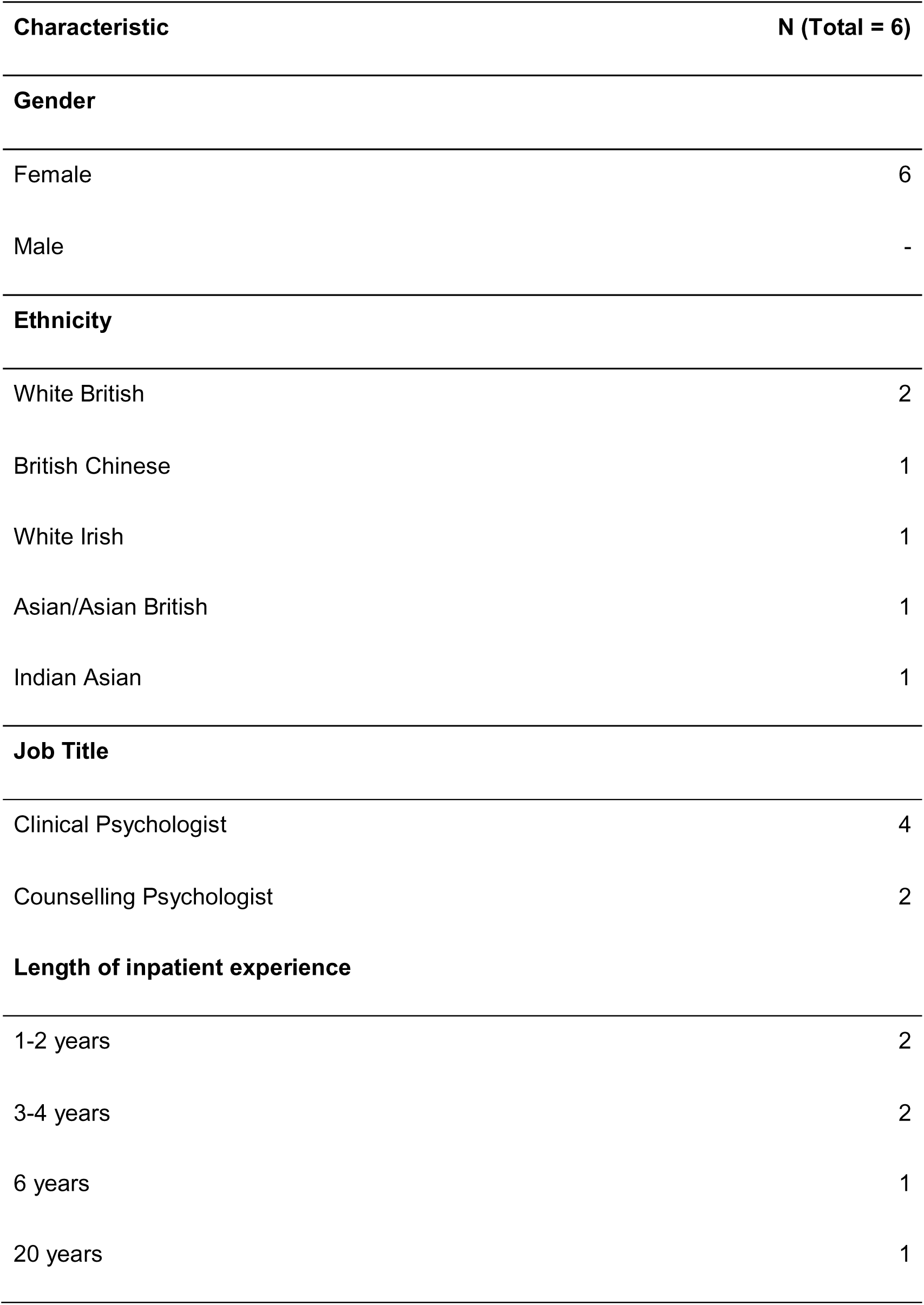
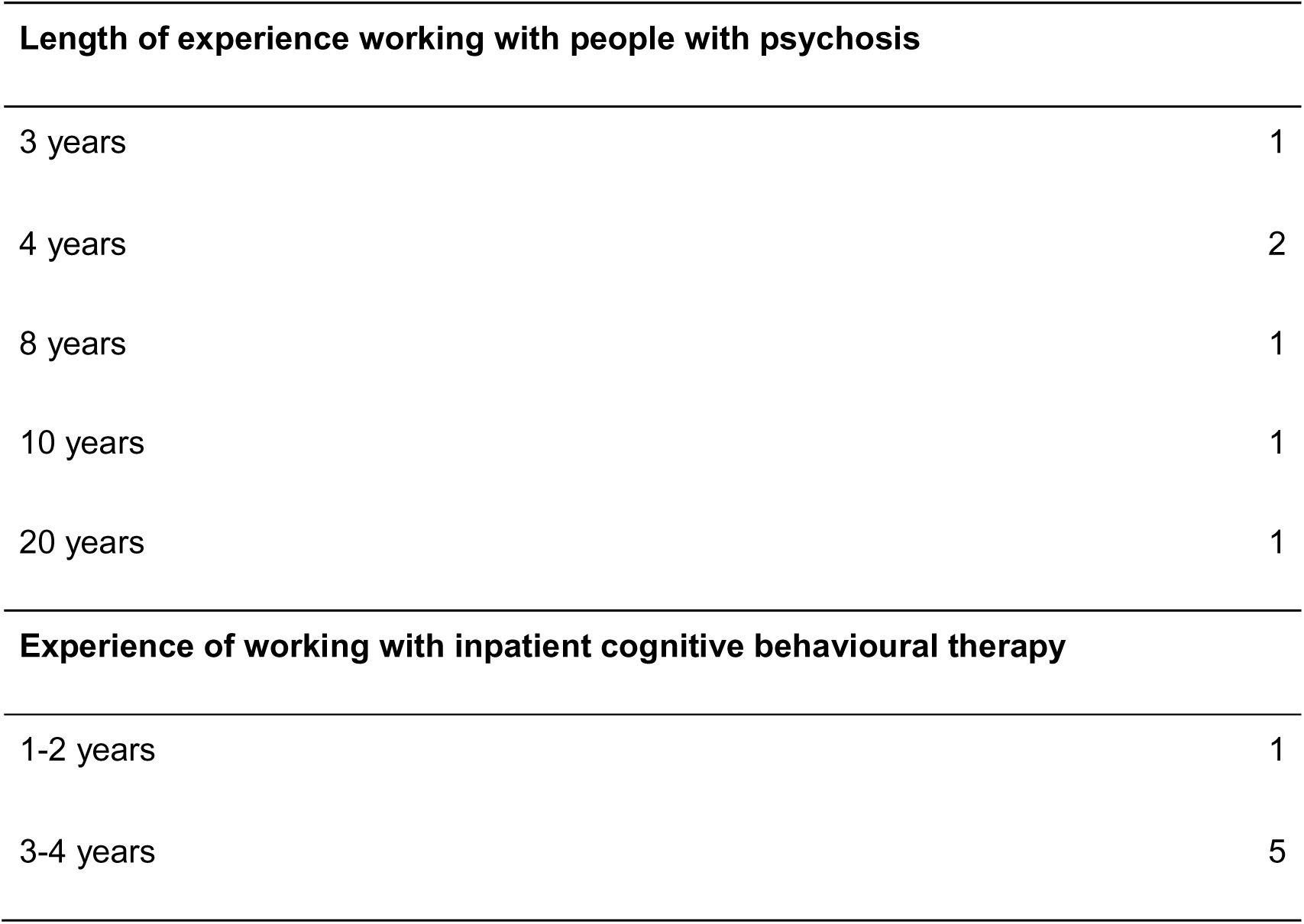
Therapist Demographics.

| **Characteristic** | **N (Total = 6)** |
| --- | --- |
| **Gender** |  |
| Female | 6 |
| Male | - |
| **Ethnicity** |  |
| White British | 2 |
| British Chinese | 1 |
| White Irish | 1 |
| Asian/Asian British | 1 |
| Indian Asian | 1 |
| **Job Title** |  |
| Clinical Psychologist | 4 |
| Counselling Psychologist | 2 |
| **Length of inpatient experience** |  |
| 1-2 years | 2 |
| 3-4 years | 2 |
| 6 years | 1 |
| 20 years | 1 |

| Length of experience working with people with psychosis |  |
| --- | --- |
| 3 years | 1 |
| 4 years | 2 |
| 8 years | 1 |
| 10 years | 1 |
| 20 years | 1 |
| Experience of working with inpatient cognitive behavioural therapy |  |
| 1-2 years | 1 |
| 3-4 years | 5 |

### Therapy log data

Session data can be found in table 3. 26 participants were randomised to CBTp+TAU, however, four participants did not participate due to moving out of the area (n=2), withdrawing, or being uncontactable after randomisation (n=2). The average number of therapy sessions delivered to the 22 participants was 7.45 (range 1-19), with an average of 4.18 sessions being missed or declined by participants. The average duration of therapy sessions conducted by the therapists was 48.27 minutes, with 60% of sessions on the ward or within the hospital grounds.

**Table 3.**
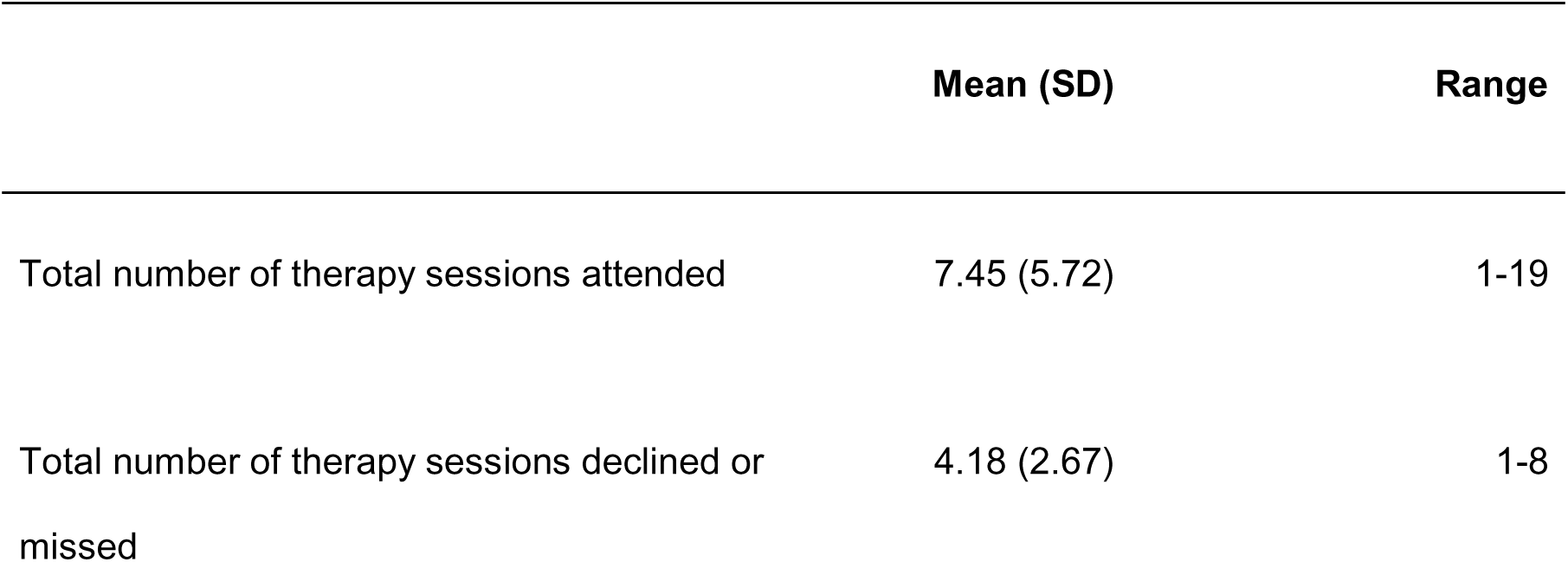
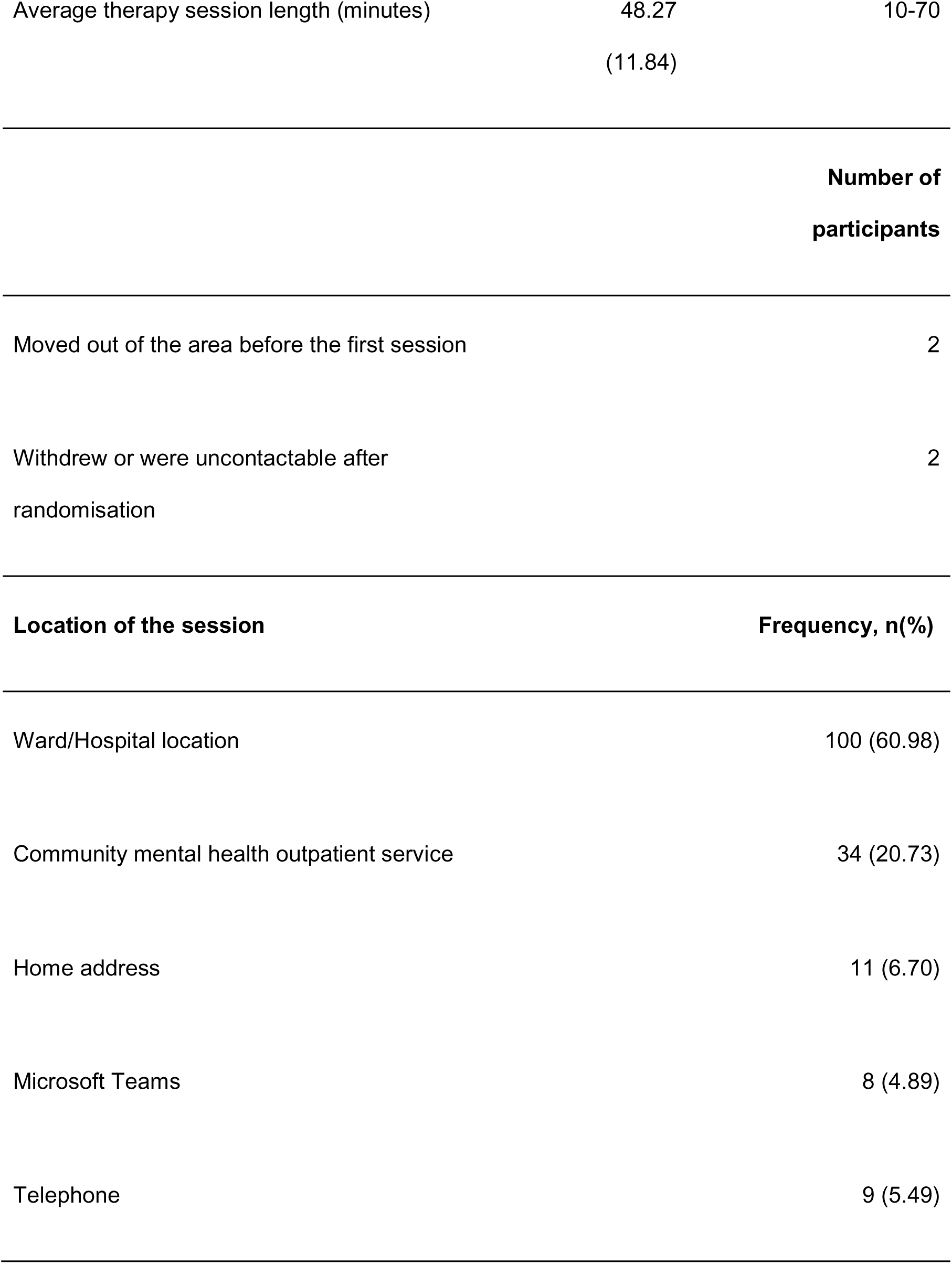
Therapy Session Attendance and Characteristics.

|  | Mean (SD) | Range |
| --- | --- | --- |
| Total number of therapy sessions attended | 7.45 (5.72) | 1-19 |
| Total number of therapy sessions declined or missed | 4.18 (2.67) | 1-8 |
| Average therapy session length (minutes) | 48.27<br>(11.84) | 10-70 |

|  |  |
| --- | --- |
|  | **Number of**<br>**participants** |

|  |  |
| --- | --- |
| Moved out of the area before the first session | 2 |
| Withdrew or were uncontactable after<br>randomisation | 2 |

|  |  |
| --- | --- |
| **Location of the session** | **Frequency, n(%)** |

|  |  |
| --- | --- |
| Ward/Hospital location | 100 (60.98) |
| Community mental health outpatient service | 34 (20.73) |
| Home address | 11 (6.70) |
| Microsoft Teams | 8 (4.89) |
| Telephone | 9 (5.49) |

Table 4 summarises the frequency of CBTp techniques used in sessions. All therapists delivered sessions to participants focused on engagement and assessment which involves a collaborative exploration of symptoms and experiences of admission. These are both considered essential components of the invention, and the remaining modules were collaboratively chosen based on the participant’s priorities. Therapists developed a formulation with 19 out of 22 (86.36%) participants aimed at understanding the links between early experiences, unhelpful thinking patterns, and current experiences and admission. Therapists delivered sessions to 16 out of 22 (72.73%) participants that included coping skills work, such as learning to manage intrusive thoughts or auditory hallucinations. Therapists supported participants in identifying and working towards various goals, for example coping with crisis, coping with voices, and staying well. Therapists were frequently involved in indirect work such as liaising with the ward team regarding care plans and liaising with secondary mental health services. In addition, therapists reported using different resources to make the sessions person-centred e.g. resources for assessment, formulation, and safety planning.

**Table 4.**
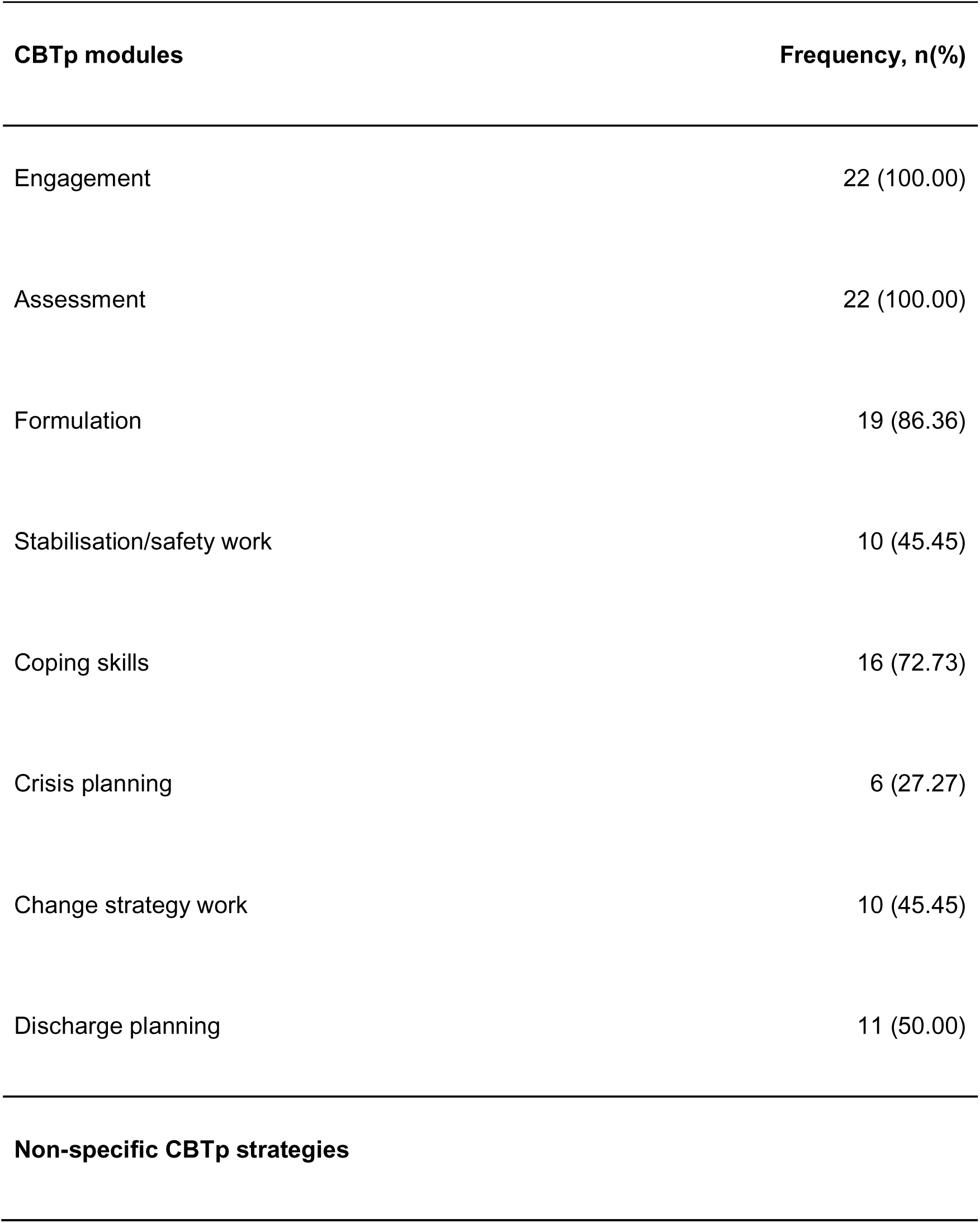
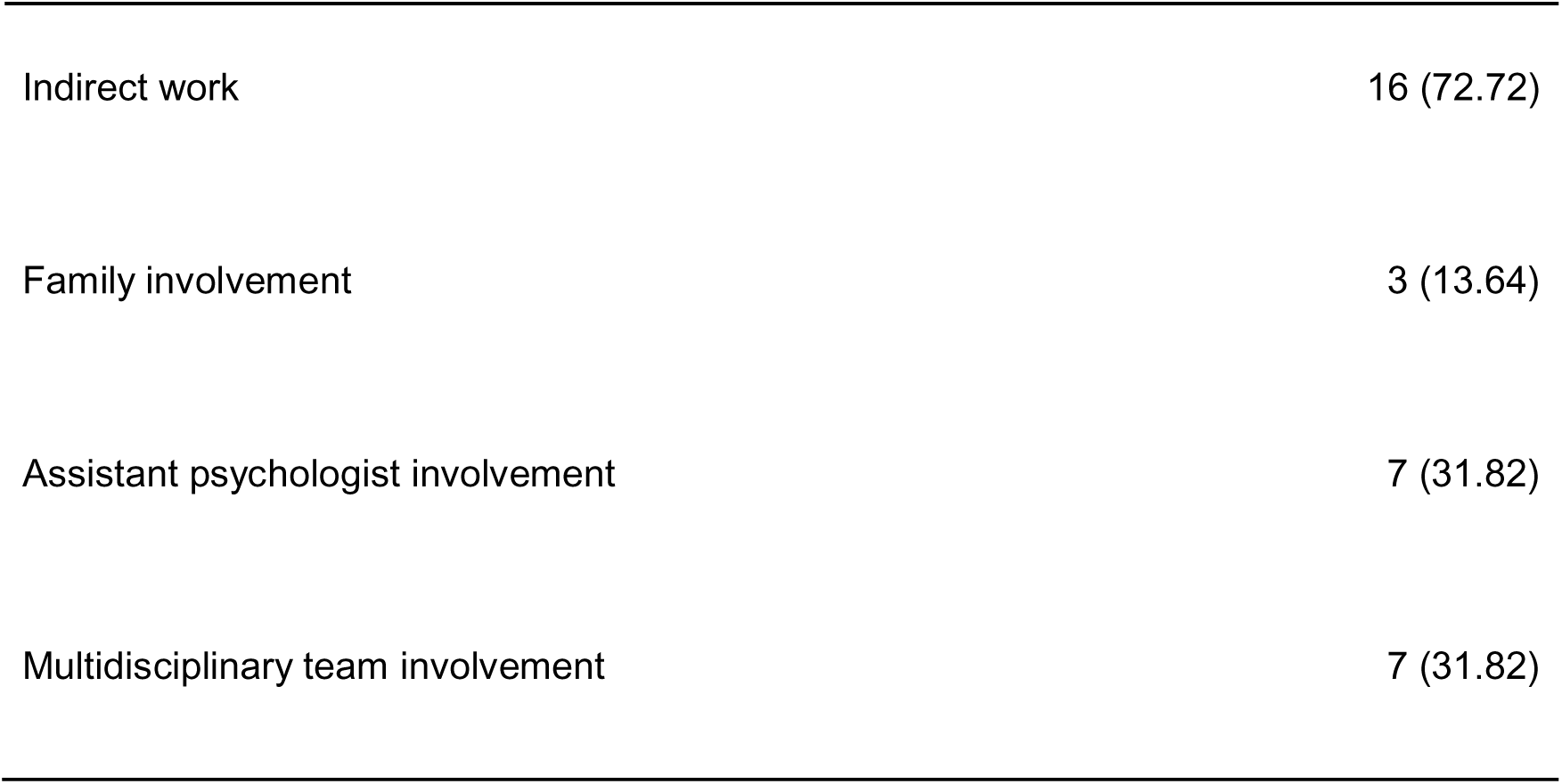
Frequency of CBTp modules used and non-specific CBTp strategies used across participants (n = total number of participants).

| **CBTp modules** | **Frequency, n(%)** |
| --- | --- |
| Engagement | 22 (100.00) |
| Assessment | 22 (100.00) |
| Formulation | 19 (86.36) |
| Stabilisation/safety work | 10 (45.45) |
| Coping skills | 16 (72.73) |
| Crisis planning | 6 (27.27) |
| Change strategy work | 10 (45.45) |
| Discharge planning | 11 (50.00) |
| **Non-specific CBTp strategies** |  |
| Indirect work | 16 (72.72) |
| Family involvement | 3 (13.64) |
| Assistant psychologist involvement | 7 (31.82) |
| Multidisciplinary team involvement | 7 (31.82) |

### Therapist interviews

A thematic analysis was undertaken on the interview data. This produced three key topic domains and related sub-themes (see Table 5). The domains were (i) therapist and patient relationship, (ii) key aspects of delivering crisis-focused therapy in inpatient settings, and (iii) challenges in delivering therapy in the acute crisis context. See Table 6 in the appendix for additional illustrative quotes.

**Table 5.** Summary of sub-themes within topic domains.

| **Therapist and patient relationship** | **Key aspects of delivering crisis-focused therapy in inpatient settings** | **Challenges in delivering therapy in the acute crisis context** |
| --- | --- | --- |
| • Facilitating safe spaces<br>• Advocacy and providing a shared understanding<br>• Empowerment, choice, and collaboration<br>• Challenges | • Adapted crisis-focused therapeutic techniques<br>• Understanding the impact of culture on psychotic experiences<br>• Flexibility in adapted CBTp model<br>• Assertive outreach<br>• Supervision and | • Disruptions of sessions due to the ward environment<br>• Navigating a short time frame<br>• Difficulty in returning to the ward when |
| discussing personal<br>experiences with<br>psychosis | training | discharged<br>• Side effects of medication |

### Domain 1: Therapist and patient relationship

This domain highlights the crucial role of the therapeutic relationship in delivering the crisis-focused CBTp-informed intervention within inpatient settings, a view that was endorsed by all therapists. Facilitating a safe therapeutic environment, which is underpinned by a trusting relationship and collaboration, was seen as essential.

### Facilitating safe spaces

Therapists emphasised the need to create a psychological and physically safe space for patients to openly express themselves and discuss their experiences of admission. They stressed the importance of focusing less on diagnosis, which is often prioritised in inpatient settings. A few therapists spoke about the importance of creating a quiet and contained environment for the session whether a private room on the ward or an outdoor space. This was instrumental in facilitating meaningful therapeutic work. Therapists found that the therapeutic relationship often had a greater impact than specific CBTp techniques, helping patients feel “validated, understood, and not judged” (Therapist 3). Key aspects of this relationship included being present, actively listening, and showing empathy which helped to foster trust and openness, allowing patients to engage in therapy.

> *“Some of it was just leaving space for him to be him and to express himself and for that not to be pathologized in quite the same way that it would be in front of the ward team.” (Therapist 1)*

### Advocacy and providing a shared understanding

All therapists highlighted the crucial role of advocacy and collaboration with the wider care team were integral to building a strong therapeutic relationship in the inpatient setting particularly considering the substantial barriers patients face. Therapists advocated for the participants and helped to address issues like inadequate living conditions or unmet support needs, ensuring a more comprehensive and supportive care approach.

Most therapists described the value of enhancing the ward team’s understanding of patients’ experiences and symptoms by presenting patient formulations during ward rounds and informal consultations. However, one therapist highlighted the challenge of facing an “ongoing view that maybe psychological intervention isn’t that helpful or isn’t a priority” (Therapist 4), noting resistance from certain members of the MDT.

> *“Thinking about the way that it incorporated things like advocacy or speaking with other members of the team, or sort of the more…the more innocuous things that you would just help someone with on the ward because they weren’t able to do it themselves or because they there were so many other barriers in place.” (Therapist 1)*

### Empowerment, choice, and collaboration

Therapists spoke about the importance of promoting patient choice and free will through collaboration, especially in the restrictive environment of an inpatient ward. A couple of therapists reflected on the inherent power dynamics in the therapeutic relationships within inpatient settings, observing that patients might feel obligated to engage due to the nature of the inpatient setting (i.e. they might be compulsorily detained against their will). To address this, therapists spoke about regularly checking in with patients and asking about their preferences to ensure that the patients had choices and felt empowered during therapy sessions.

> *“I noticed that session where I felt that he had something that had been taken in and something had been installed in his agency to feel that he had more power over their voices or he was more in control.” (Therapist 3).*

### Challenges discussing personal experiences with psychosis

Therapists noted that patients were sometimes reluctant to share their experiences of psychosis, which they assumed was due to feelings of shame and stigma. However, one therapist said that patients often expressed a strong desire to share their stories and have their experiences acknowledged, even when this involved revealing vulnerable aspects of their lives. To address these challenges, therapists implemented strategies aimed at fostering a trusting and non-judgmental therapeutic relationship. Therapists noted that this helped to alleviate feelings of shame and stigma and encouraged patient openness.

> *“It’s really vulnerable, exposing parts of people’s minds.…. Yeah, every person struggled and yet appeared to want to share their story. Have somebody witness their narrative.” (Therapist 5)*

### Domain 2: Key aspects of delivering crisis-focused therapy in inpatient settings

All therapists discussed critical factors necessary for delivering the adapted CBTp in inpatient settings. They highlighted the importance of person-centred, flexible therapeutic approaches along with robust training and supervision to effectively support the unique needs of individuals with psychosis in inpatient settings.

### Adapted crisis-focused therapeutic approaches

All therapists discussed the helpfulness of adapting therapeutic approaches for individuals with psychosis in inpatient settings. Therapists emphasised the importance of tailoring therapeutic techniques to address the reasons for the patient’s admission, helping them gain insight into their psychosis and the contributing factors. This approach was novel and beneficial for many patients who had not previously received psychological interventions. Therapists described how adapting therapeutic approaches such as formulation and assessment, helped mitigate the impact of psychotic symptoms like hearing voices. For example, one therapist spoke about how the intervention enhanced patients’ coping skills by enabling them to challenge negative thoughts related to the voices.

Consequently, the patient began to reinterpret distressing psychotic experiences, leading to a reduction in distress.

> *“When we did a bit of the psychoeducation around psychosis, so her being able to make sense of her experience or what was happening to her, which probably felt extremely confusing for a young person….I’m arguing definitely helped because there was definitely a shift in her way of being with me in future interactions.” (Therapist 6)*

### Understanding the Impact of Culture on Psychotic Experiences

Most therapists emphasised the importance of cultural awareness in therapy, highlighting the need to consider patients’ cultural backgrounds, values, and beliefs to better understand their experiences and tailor therapeutic approaches. One therapist spoke about actively discussing how cultural beliefs and experiences can influence a patient’s early life and their experiences of psychosis. This included exploring how culture affected both the manifestation and interpretation of psychotic symptoms and diagnoses, ensuring more comprehensive and personalised therapeutic interventions.

Another therapist highlighted the importance of addressing racial perceptions in therapy. They discussed how they incorporated the client’s experiences and views on race into the therapeutic formulation, particularly given that the client was working with a predominantly white team and was new to Westernised therapy approaches. This required careful consideration of how cultural differences, including the therapist’s background, impacted the therapeutic relationship and the client’s understanding of therapy.

> *“We definitely spoke about race actually. I only had clients which were not white. One of them again, we spoke about kind of perceptions and racial perceptions and that was his formulation. Being able to name that and explain that and also talk about this being a white woman across the team and from him talking about Westernised therapy when he’s not quite sure what it is and that was his first experience of psychology.” (Therapist 5)*

### Flexibility in the adapted CBTp model

Therapists highlighted the importance of flexibility in the inpatient environment, such as holding sessions outdoors or in patients’ rooms which was essential due to the lack of quiet spaces on the ward. This flexibility also extended to adjusting session frequency and duration to accommodate patients’ preferences and conflicting schedules, which helped to navigate the constraints of the environment and maintain patient engagement.

> *“This particular adapted model of CBTp in my view offers all of that flexibility because there was never any pressure on me, as I see as a person delivering CBT for psychosis…there’s no pressure on me to do it in a very particular way.” (Therapist 6)*

### Assertive outreach

All therapists spoke about appreciating the adapted CBTp model for its emphasis on proactive engagement and assertive outreach with patients who might otherwise miss sessions. This approach involved actively reaching out to patients to ensure they attended their appointments, addressing barriers to attendance, and offering alternative methods to engage them in therapy. Most therapists highlighted the importance of flexibility in communication methods, such as using text messages instead of letters or phone calls. This adjustment helped in effectively reaching patients, particularly those who found conventional methods less reliable or engaging. Therapists also noted that the protocol allowed them to provide multiple opportunities for patients to continue therapy even after missed sessions, which contrasted with community settings where discharge often followed missed appointments.

> *“The people that did stay in therapy, they did miss a lot of sessions with me at various points. So I think maybe if I’d have been part of a sort of typical Community therapy team, they would have been discharged. So I suppose that’s a good example of what…might maintain people’s engagement if it can be a bit more flexible and not just discharge them after they’ve missed a couple of sessions.” (Therapist 4)*

### Supervision and Training

Therapists highlighted the value of supervision for its flexibility, adaptability, and effectiveness in providing support and helping to address the challenges they faced.

Therapists spoke about how supervision provided essential support for troubleshooting and developing therapeutic techniques and reflected the flexible approach of the adapted CBTp model itself. In addition, therapists found the training to be thorough and highly valuable, offering both essential theoretical knowledge and practical skills for delivering the adapted CBTp in inpatient settings.

> *“I think it [training] was really useful and it was helpful. Yeah, I remember it helped me to see how flexible the model could be and also sort of applying the model for use in crisis as well.” (Therapist 4)*

### Domain 3: Challenges in delivering therapy in the acute crisis context

Therapists highlighted several challenges in delivering therapy within the acute crisis context. These challenges included practical issues related to the ward environment as well as practical difficulties such as navigating the brief intervention time frame.

### Disruption of sessions due to ward environment

One of the key challenges in the delivery of the intervention whilst on the ward was finding a quiet space without interruptions. All therapists highlighted barriers such as constant interruption, noise levels, and high acuity within the ward setting. These factors impacted the therapeutic process affecting session structure, patient’s mood, and at times the ability to create a safe, conducive environment for effective therapy.

> *“It doesn’t always feel like a safe therapeutic environment because…there might be people outside your rooms, you might get kicked out of your rooms and that kind of thing does impact on it.” (Therapist 4)*

### Navigating a short time frame

One therapist discussed initially finding it challenging to balance the intensive needs of inpatient care within the brief intervention timeframes. They noted challenges in structuring sessions effectively while ensuring enough time to build a therapeutic relationship, address complex needs, and thoroughly assess and formulate the patient’s history and current experiences. The therapist spoke about how their training equipped them with strategies to overcome these challenges, enabling them to adapt session content to remain person-centred while fitting within the constrained time frames.

> *“I think initially with the first kind of time that I did it was a little bit difficult to kind follow that kind of structure and not to spend too much time on things that maybe wasn’t so relevant. But still getting enough kind of like history or, knowledge around what led them to their crisis and the psychosis, psychotic experience, things like that”. (Therapist 2)*

### Difficulty in returning to the ward when discharged

Many therapists spoke of challenges in maintaining engagement with patients after they were discharged from the hospital. Therapists believed that many patients found returning to the ward for therapy sessions difficult due to their negative experiences in the hospital or their associations with the hospital with their crisis. To overcome challenges in maintaining engagement, therapists adopted a flexible, patient-centred approach and where possible offered alternative locations for therapy sessions e.g. community mental health settings or remote sessions which helped patients to engage in therapy.

> *“I think one of the difficulties is that when they left hospital them coming back and to see me and have the sessions were actually quite traumatic because of the hospital experience that they had and so that was difficult to kind of keep that engagement.” (Therapist 2)*

### Side effects of medication

Therapists noted that in inpatient settings, patients are often on higher doses of psychiatric medications compared with community mental health settings. This resulted in significant medication side effects, particularly drowsiness and cognitive impairment which hindered patients’ engagement with therapy sessions. Therapists supported patients by revisiting and summarising key therapeutic interventions in multiple sessions to ensure they had a clear understanding.

> *“Often there are sort of high levels of medication being used, so I know for at least two of the service users that I worked with, they would often be quite drowsy when I visited them on the ward.” (Therapist 4)*

## Discussion

This study aimed to examine the acceptability of delivering crisis-focused CBTp-informed intervention in inpatient settings from the therapist’s perspective.

Therapists delivered an average of 7.45 sessions per patient (range 1-19), aligning with the adapted CBTp model of six to eight sessions. Continuity was often facilitated by therapists offering additional sessions in community settings after patients were discharged from the ward. The average number of sessions declined or missed was 4.18 and these were due to, for example, patients not feeling well, being on leave from the ward, having other conflicting appointments, or being uncontactable. This highlights the importance of therapists remaining flexible and adaptable when working in an inpatient setting, which is supported by a recent Delphi study that found therapists delivering CBTp in inpatient settings need to be more flexible with session content and delivery (Wood et al., 2022a). In the qualitative interviews, therapists reported that flexibility and adaptability were crucial due to the high levels of distress experienced by patients and the constraints of the inpatient environment. These insights suggest that, to maintain continuity of care and patient engagement, therapists may need to make allowances for missed sessions, reschedule appointments, and adjust both the location and agenda of sessions as needed.

The data from the therapy log shows that after the initial engagement and assessment sessions that all therapists offered to patients, most therapists collaboratively developed a formulation with patients aimed at better understanding their experiences and current admissions. These modules were essential, so it was expected that these should be offered to all participants however some patients only had one therapy session which may have made the formulation more challenging to complete. In addition, the results from the qualitative interviews found that therapists emphasised the importance of adapting crisis-focused therapeutic approaches, such as assessment and formulation, to help patients understand their current crisis and the reasons for their admission. Therapists reflected that this facilitated empowerment and supported patients to work towards their goals and enabled effective relapse prevention and discharge planning. This approach aligns with previous research showing that crisis-planning interventions can reduce the risk of compulsory hospital admissions by 25% for individuals with psychotic illness or bipolar disorder (Molyneaux et al., 2019).

Also, therapists were guided by the recovery goals set by the patients, which included coping with crisis and relapse prevention. This approach is supported by previous research, which demonstrated that self-management interventions such as psychoeducation and relapse prevention led to improvements for individuals diagnosed with serious mental illness (Lean et al., 2019). Additionally, indirect work played a significant role in the delivery of crisis-focused CBTp in inpatient settings, with therapists engaging in indirect work for 73% of patients, such as liaising with the ward team and attending professional meetings.

Previous research has shown that support and involvement of MDT within an inpatient setting are essential for maximising the effectiveness of psychological therapy (Wood et al., 2019). These findings suggest that therapists should deliver an adapted crisis-focused patient-centred CBTp model in inpatient settings, prioritising tailored self-management interventions while ensuring collaboration with the MDT both on the ward and in the community.

The qualitative interviews also uncovered key factors for delivering the therapy that were not highlighted in the therapy log data. Therapists emphasised the importance of the therapeutic relationship when delivering the adapted CBTp-informed intervention. They highlighted the need to create a safe therapeutic environment where patients can openly share their experiences of psychosis and their admission, ensuring they feel validated and understood. The significance of a strong therapeutic relationship has been highlighted in other qualitative evaluations of CBTp (Berry & Hayward, 2011; Wood et al., 2015). We identified that advocacy and collaboration with the broader care team, extending beyond traditional CBTp, are crucial for cohesive teamwork and a unified understanding of patient needs and psychological formulations. Importantly, the patients who took part in the main CRISIS study, i.e. those who received therapy from the therapists discussed in this paper, also recognised the importance of a strong therapeutic relationship, characterised by advocacy, active listening, and empathy (Wood et al., 2024b). Therapists also acknowledged the inherent power imbalances within inpatient settings that can affect the patient-therapist relationship and influence the therapeutic alliance. Therefore, therapists should be mindful of these dynamics when delivering CBTp in inpatient settings, as such imbalances may be amplified in these environments (Morgan et al., 2023).

Another important aspect of delivering the therapy was recognising the impact of culture on psychotic experiences, which is not commonly incorporated into traditional CBTp practices. In inpatient settings, where patients from black and ethnic minority backgrounds are overrepresented (NHS Digital, 2024). Therapists noted that openly discussing potential cultural influences during therapy sessions was beneficial for patients. Therefore, therapists must consider the intersectionality of patients’ identifies and be attuned to culturally specific social graces, which shape patients’ experiences. Therapists should actively engage in conversations about patients’ cultural backgrounds, rather than avoiding these discussions (Naeem et al., 2019). The coproduction group involved in the CRISIS study also emphasised the importance of therapists seeking to understand a patient’s individual cultural, religious, and spiritual beliefs as well as inquiring about any negative experiences including racism (Wood et al., 2023).

Finally, all therapists identified several challenges in delivering the adapted CBTp in the acute crisis context. Practical barriers such as high acuity of the ward environment, nosiness, and lack of room space were highlighted by therapists, which has also been highlighted in previous research (Jacobsen, 2019; Wood et al., 2019). One therapist highlighted the difficulty of maintaining trusting relationships while conducting thorough assessments within a limited time frame, noting that effective training and supervision were essential for managing these constraints. To address these challenges, regular and flexible training and supervision are recommended.

Our research found that the training of the CBTp-informed crisis intervention and its delivery was acceptable to therapists. Acceptability was assessed based on therapists’ perceptions of the intervention’s feasibility, effectiveness, and overall satisfaction with its delivery. This approach aligns with the Sekhon framework for acceptability (Sekhon et al., 2017), which identifies perceived effectiveness and ease of implementation as key factors in determining acceptability from the perspective of those delivering the intervention. Our results show that a further large-scale RCT is needed to determine the efficacy of the intervention. Therefore, future research should focus on a full-scale effectiveness trial of the adapted CBTp in inpatient settings.

It is important to consider both the strengths and limitations of this research. A key strength is that this research offers new insights into how therapists delivered a crisis-focused CBTp-informed intervention and their perspectives on its delivery and training in culturally diverse and acute inpatient setting, an area that has not been previously explored. The therapy-specific data log provides comprehensive documentation of therapy sessions which enhances the understanding of the therapy process and delivery and allows for replication of the therapy in future research trials and clinical practice. By combining the therapy log with qualitative data, this study provides a richer understanding of both the structured delivery of the intervention and therapists’ real-world experiences. This approach allows for a more nuanced evaluation, helping to refine and adapt the therapy to better meet the needs of patients in diverse inpatient settings.

Several limitations were identified for this study. For instance, the therapy log may not capture broader contextual factors, such as changes in the ward environment or shifts in patient circumstances, which can influence therapy delivery. This limitation is addressed through follow qualitative interviews but we did not explicitly ask why participants did not deliver certain aspects of the intervention, which was a limitation. The involvement of LW in supervising this research raises concerns about conflict of interest. To address this concern, AR led all aspects of the analysis, and the interviews were conducted by independent research assistants who were not associated with the trial. All interviews were conducted remotely, which may have impacted the richness of the data (Johnson et al., 2021), however, some researchers argue that remote interviews can yield data as rich as in-person interviews (Abrams et al., 2015).

In conclusion, the delivery of a crisis-focused CBTp-informed intervention and relevant training for the therapists, was found to be acceptable within inpatient settings from the perspectives of the therapists delivering it. This study has been able to identify the potential key beneficial components of the intervention and key barriers and facilitators of the intervention specific to these settings.

## Data Availability

All data produced in the present study are available upon reasonable request to the authors

## Appendix

**Table 6.** Additional illustrative quotes.

| Therapist and patient relationship |  |
| --- | --- |
| **1.1 Facilitating safe spaces** | “I think it [therapy] was helpful. I think a lot was about ... having grounding strategies and kind of more self-soothing things and psychoeducation..... and actually looking at components that will help them recover in the sense of what they wanted to achieve and their goals.” (Therapist 2) |
| **1.2 Advocacy and providing a shared understanding** | *“I was also acting as an advocate and would sort of bring his formulation repeatedly to ward rounds and that kind of thing, then to try and enable him to be understood and to create less confusion between staff members as well. Which had a knock-on effect in his care.” (Therapist 1).* |
| **1.3 Empowerment, choice and** | *“I think sometimes he would be a bit compliant with me ... the power dynamics of the ward that generally staff members are* |
| **collaboration** | *telling the patients what to do and that there might be a perception of consequence if that doesn't happen." (Therapist 1)* |
| **2.1 Key aspects of delivering crisis-focused therapy in inpatient settings** |  |
| **2.1 Adapted crisis-focused therapeutic approaches** | *"When we did a bit of the psychoeducation around psychosis, so her being able to make sense of her experience or what was happening to her, which probably felt extremely confusing for a young person. I think definitely... I'm arguing definitely helped because there was definitely a shift in her way of being with me in future interactions." (Therapist 6)* |
| **2.2 Understanding the Impact of Culture on Psychotic Experiences** | *"I think one of the things that we did look at, as part of the CBTp templates that we did, was kind of include cultural aspects and actually thinking about how someone's culture impacts either their early experiences or their experiences of having a psychosis diagnosis." (Therapist 2)* |
| **2.3 Flexibility in the adapted CBTp model** | *"Sometimes it [barriers to delivering therapy] was just other things that they had got, that social stuff where they couldn't make it. Other focuses, sometimes they would kind of, yeah maybe have a different focus because something had happened in their week and wanted some spend some time, kind of maybe talking about that which was which we did". (Therapist 2)* |
|  | *"I'm just thinking of my patient that had a lot of immigration issues. I think maybe some of the more direct CBTp techniques ..... like coping skills were probably less well received. The social situation was so challenging. ...I know this this model of CBTp is meant to sort of be inclusive and it include things like that which you think it does. But I just think for some people I guess direct CBTp sort of skills might not be their main kind of treatment need. It might be more about thinking about their wider life and the context that they're in." (Therapist 4)* |
| **2.4 Assertive outreach** | *"My other clients I would say yeah, on average probably like DNA and every other session. So I guess struggling to kind of reliably get to their appointments and I found what I found helpful actually was, I guess, doing, prompting, sending texts." (Therapist 4)* |
| **2.5 Supervision and Training** | *"Supervision was excellent... Is very flexible, very adaptable and mirrored the therapy itself." (Therapist 5)* |
| **3. Challenges in delivering therapy in the acute crisis context** |  |
| **3.1 Disruption of sessions due to ward environment** | *"On the ward, availability would be a problem, actually, I guess if you're delivering therapy quite flexibly and also people aren't really working out meeting rooms and that kind of thing. The room was sometimes not available. I mean, people would also* |
|  | *walk in all the time. “ (Therapist 1).* |
| **3.2 Navigating a short time frame** | *“Difficult sometimes to focus and on a piece of work within the kind of the limited time. Umm...but again, I think just having that kind of focus on what was the experience and really just focused in what they wanted to work on and having a collaborative goal help us kind of keep track of what the aim of the of the study and the aim of the research in the therapy I guess.” (Therapist 2)* |
|  | *“It's difficult for someone, for example, to work on understanding the voices and actually being quite anxious about talking about their experiences or in the in that moment they were experiencing that and feeling quite overwhelmed. I guess it wasn't a massive barrier, but sometimes that could have. It really was important for the work to be a little bit slower at times and to continue and to kind of check in with the with the patient and how they are, whether they wanted to continue talking about a certain thing or having more tools for them to manage and cope.” (Therapist 2)* |

**Table 7.**
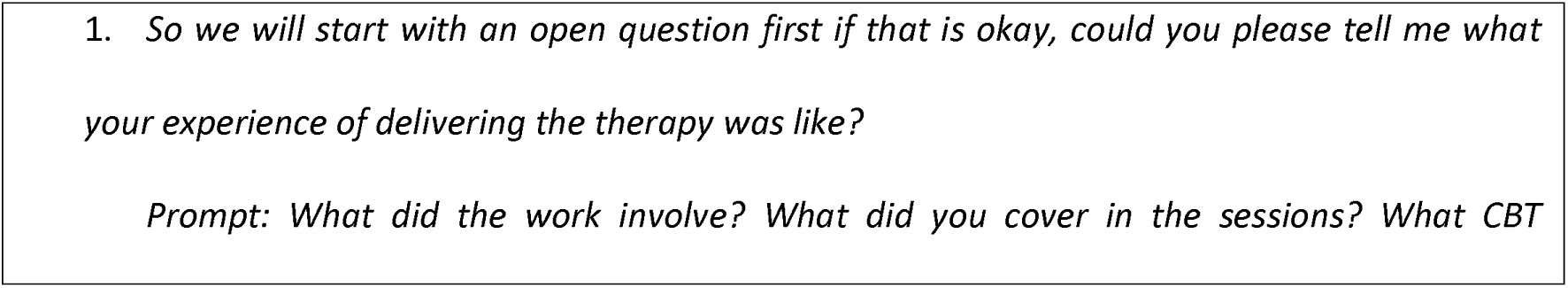
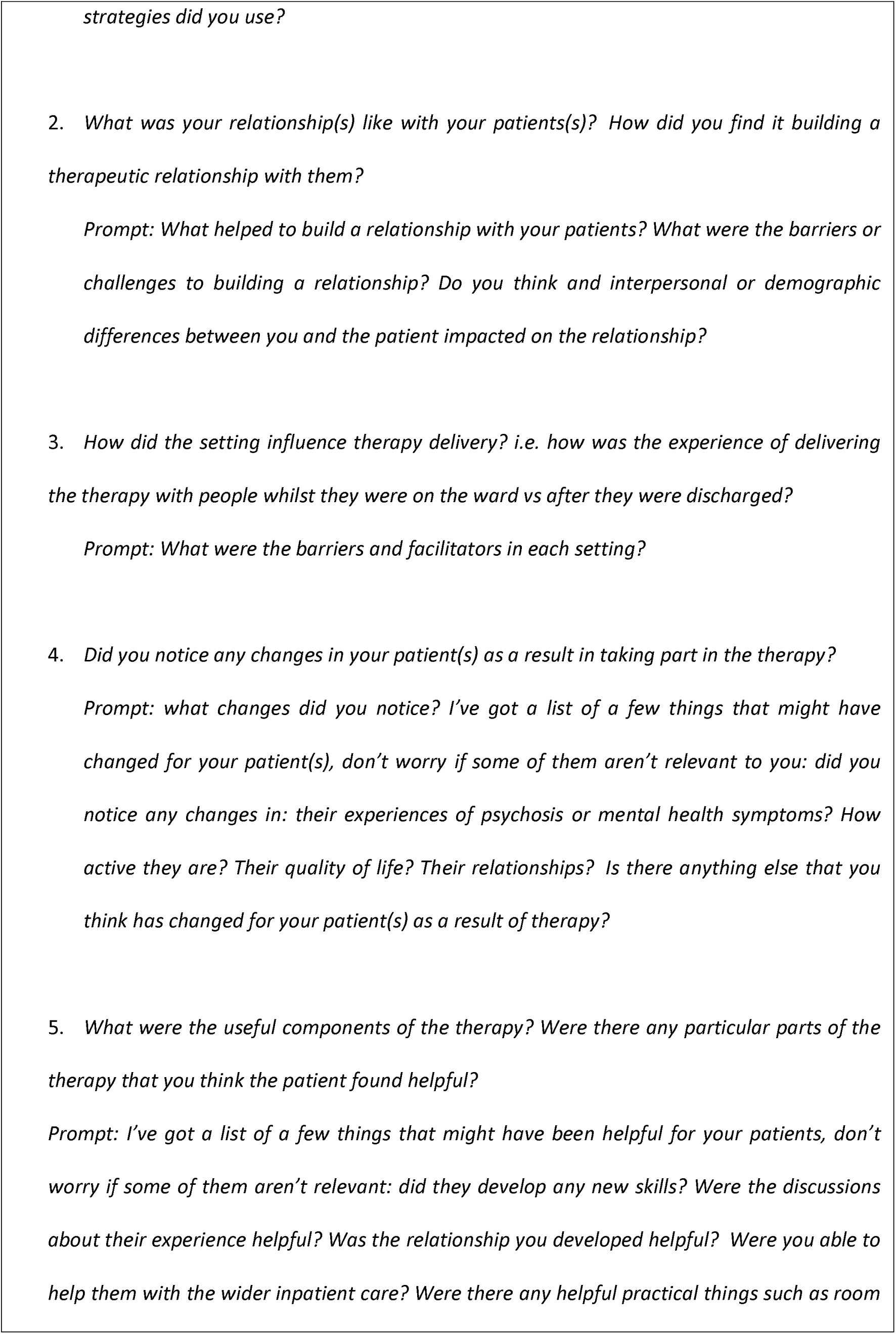
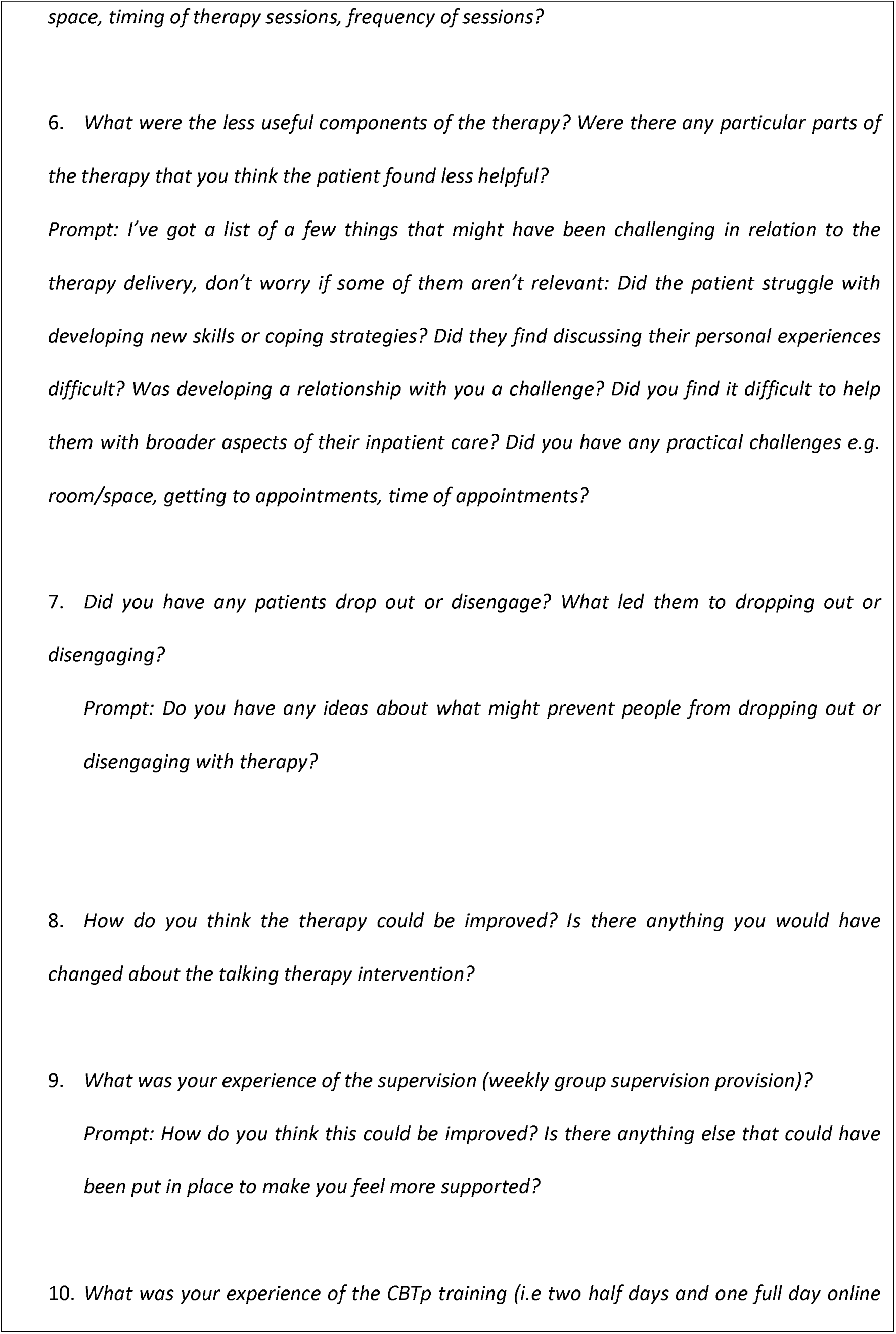
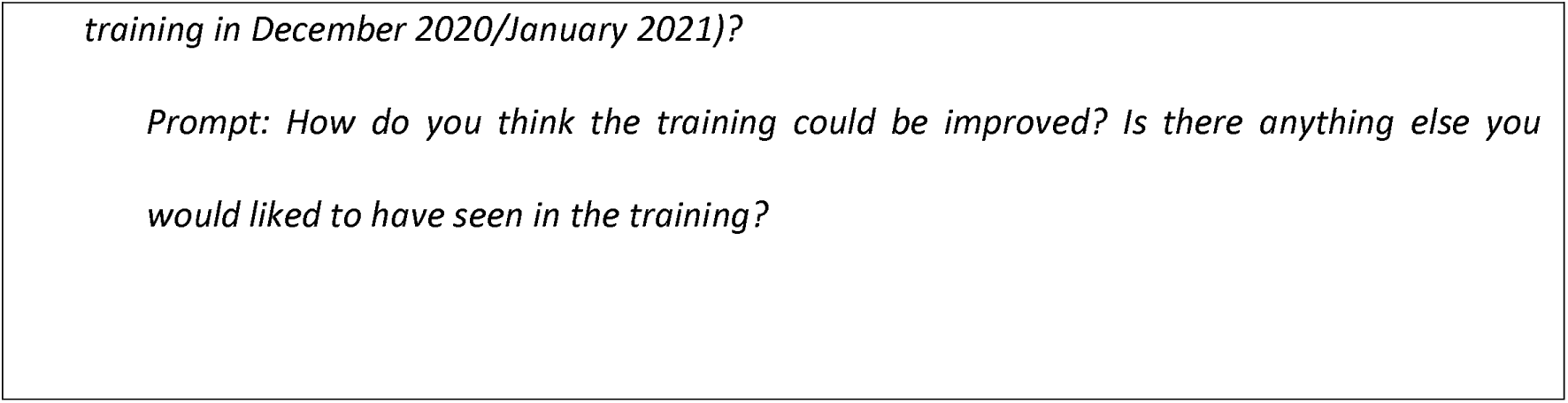
List of semi structured interview questions.

## References

Abrams, K. M., Wang, Z., Song, Y. J., & Galindo-Gonzalez, S. (2015). Data richness trade-offs between face-to-face, online audiovisual, and online text-only focus groups. Social Science computer review, 33(1), 80–96.

Berry, C., & Hayward, M. (2011). What can qualitative research tell us about service user perspectives of CBT for psychosis? A synthesis of current evidence. Behavioural and Cognitive Psychotherapy, 39(4), 487–494.

Berry, K., Raphael, J., Haddock, G., Bucci, S., Price, O., Lovell, K., Drake, R. J., Clayton, J., Penn, G., & Edge, D. (2022). Exploring how to improve access to psychological therapies on acute mental health wards from the perspectives of patients, families, and mental health staff: qualitative study. BJPsych Open, 8(4). 10.1192/bjo.2022.513

Blackburn, I. M., James, I. A., Milne, D. L., Baker, C., Standart, S., Garland, A., & Reichelt, F. K. (2001). The revised cognitive therapy scale (CTS-R): psychometric properties. Behavioural and cognitive psychotherapy, 29(4), 431–446.

Bowers, L. (2014). Safewards: a new model of conflict and containment on psychiatric wards. Journal of psychiatric and mental health nursing, 21(6), 499–508.

Braun, V., & Clarke, V. (2006). Using thematic analysis in psychology. Qualitative research in psychology, 3(2), 77–101.

Care Quality Commission. (2009). Supporting briefing note; issues highlighted by 2009 survey of mental health acute inpatient services. Newcastle upon Tyne: Care Quality Commission.

Evlat, G., Wood, L., & Glover, N. (2021). A systematic review of the implementation of psychological therapies in acute mental health inpatient settings. Clinical Psychology & Psychotherapy, 28(6), 1574–1586.

Halvorsrud, K., Nazroo, J., Otis, M., Brown Hajdukova, E., & Bhui, K. (2018). Ethnic inequalities and pathways to care in psychosis in England: a systematic review and meta-analysis. BMC medicine, 16, 1–17.

Harris, R., Maguire, T., & Newman-Taylor, K. (2022). What do trainee cognitive behavioural therapists need from clinical supervision to develop their skills in working with people with psychosis? A qualitative analysis. Psychosis, 14(2), 120–130.

Hoffmann, T. C., Glasziou, P. P., Boutron, I., Milne, R., Perera, R., Moher, D., Altman, D. G., Barbour, V., Macdonald, H., Johnston, M., Lamb, S. E., Dixon-Woods, M., McCulloch, P., Wyatt, J. C., Chan, A.-W., & Michie, S. (2014). Better Reporting of interventions: Template for Intervention Description and Replication (TIDieR) Checklist and Guide. BMJ, 348, g1687–g1687. 10.1136/bmj.g1687

Jacobsen, P., Hodkinson, K., Peters, E., & Chadwick, P. (2018). A systematic scoping review of psychological therapies for psychosis within acute psychiatric in-patient settings. The British Journal of Psychiatry, 213(2), 490–497.

Johnson, D. R., Scheitle, C. P., & Ecklund, E. H. (2021). Beyond the in-person interview? How interview quality varies across in-person, telephone, and Skype interviews. Social science computer review, 39(6), 1142–1158.

Kings Fund. (2024). Mental health 360: acute mental health care for adults. https://www.kingsfund.org.uk/insight-and-analysis/long-reads/mental-health-360-acute-mental-health-care-adults

Lean, M., Fornells-Ambrojo, M., Milton, A., Lloyd-Evans, B., Harrison-Stewart, B., Yesufu-Udechuku, A., Kendall, T., & Johnson, S. (2019). Self-management interventions for people with severe mental illness: Systematic review and meta-analysis. The British Journal of Psychiatry, 214(05), 260–268. 10.1192/bjp.2019.54

McKenna, P., & Kingdon, D. (2014). Has cognitive behavioural therapy for psychosis been oversold?. Bmj, 348.

Molyneaux, E., Turner, A., Candy, B., Landau, S., Johnson, S., & Lloyd-Evans, B. (2019). Crisis-planning interventions for people with psychotic illness or bipolar disorder: systematic review and meta-analyses. BJPsych open, 5(4), e53. 10.1192/bjo.2019.28

Morgan, C., Clarkson, L., Hiscocks, R., Hopkins, I., Berry, K., Tyler, N., Wood, L., & Jacobsen, P. (2023). What should inpatient psychological therapies be for? Qualitative views of service users on outcomes. Health Expectations. 10.1111/hex.13889

Naeem, F., Phiri, P., Rathod, S., & Ayub, M. (2019). Cultural adaptation of cognitive– behavioural therapy. BJPsych advances, 25(6), 387–395.

National Institute for Health and Care Excellence (2014). Psychosis and schizophrenia: treatment and management.

NHS Benchmarking. (2016). 2016 benchmarking of adult and older people’s mental health services.

NHS Digital. (2024). Mental Health Bulletin, 2022-23 Annual report. NHS Digital. https://digital.nhs.uk/data-and-information/publications/statistical/mental-health-bulletin/2022-23-annual-report

NHS England. (2016). The Five Year Forward View for Mental Health. In https://www.england.nhs.uk/wp-content/uploads/2016/02/Mental-Health-Taskforce-FYFV-final.pdf.

NHS England. (2019). The NHS Long Term Plan. NHS Long Term Plan. https://www.longtermplan.nhs.uk/publication/nhs-long-term-plan/

NHS England. (2023). “*Acute Inpatient Care for Adults and Older Adults*.” NHS England.

Paterson, C., Karatzias, T., Dickson, A., Harper, S., Dougall, N., & Hutton, P. (2018). Psychological therapy for inpatients receiving acute mental health care: A systematic review and meta-analysis of controlled trials. British Journal of Clinical Psychology, 57(4), 453–472.

Qassem T, Bebbington P, Spiers N, McManus S, Jenkins R, Dein S. Prevalence of psychosis in black ethnic minorities in Britain: analysis based on three national surveys. Soc Psychiatry Psychiatr Epidemiol. 2015 Jul;50(7):1057–64. doi: 10.1007/s00127-014-0960-7. Epub 2014 Sep 11. PMID: 25208909; PMCID: PMC4464643.

Radcliffe, J., & Bird, L. (2016). Talking therapy groups on acute psychiatric wards: patients’ experience of two structured group formats. BJPsych bulletin, 40(4), 187–191.

Rathod, S., Phiri, P., Harris, S., Underwood, C., Thagadur, M., Padmanabi, U., & Kingdon, D. (2013). Cognitive behaviour therapy for psychosis can be adapted for minority ethnic groups: a randomised controlled trial. Schizophrenia research, 143(2-3), 319–326.

Royal College of Psychiatrists. (2015). Standards for inpatient mental health services.

Schlief, M., Rich, N., Rains, L. S., Baldwin, H., Rojas-Garcia, A., Nyikavaranda, P., Persaud, K., Dare, C., French, P., Lloyd-Evans, B., Crawford, M., Smith, J., Kirkbride, J. B., & Johnson, S. (2023). Ethnic differences in receipt of psychological interventions in Early Intervention in Psychosis services in England – a cross-sectional study. Psychiatry Research, 115529. 10.1016/j.psychres.2023.115529

Sekhon, M., Cartwright, M., & Francis, J. J. (2017). Acceptability of healthcare interventions: an overview of reviews and development of a theoretical framework. BMC health services research, 17, 1–13.

Skivington, K., Matthews, L., Simpson, S. A., Craig, P., Baird, J., Blazeby, J. M., Boyd, K. A., Craig, N., French, D. P., McIntosh, E., Petticrew, M., Rycroft-Malone, J., White, M., & Moore, L. (2021). A New Framework for Developing and Evaluating Complex interventions: Update of Medical Research Council Guidance. BMJ, 374(1), n2061. 10.1136/bmj.n2061

Small, C., Pistrang, N., Huddy, V., & Williams, C. (2018). Individual psychological therapy in an acute inpatient setting: Service user and psychologist perspectives. Psychology and Psychotherapy: Theory, Research and Practice, 91(4), 417–433.

Tong, A., Sainsbury, P., & Craig, J. (2007). Consolidated criteria for reporting qualitative research (COREQ): a 32-item checklist for interviews and focus groups. International journal for quality in health care, 19(6), 349–357.

Voskes, Y., Van Melle, A. L., Widdershoven, G. A., van Mierlo, A. F. M. M., Bovenberg, F. J., & Mulder, C. L. (2021). High and intensive care in psychiatry: a new model for acute inpatient care. Psychiatric Services, 72(4), 475–477.

Wood, L., Burke, E., & Morrison, A. (2015). Individual cognitive behavioural therapy for psychosis (CBTp): a systematic review of qualitative literature. Behavioural and cognitive psychotherapy, 43(3), 285–297.

Wood, L., Williams, C., Billings, J., & Johnson, S. (2019). Psychologists’ perspectives on the implementation of psychological therapy for psychosis in the acute psychiatric inpatient setting. Qualitative Health Research, 29(14), 2048–2056.

Wood, L., Williams, C., Billings, J., & Johnson, S. (2020). A systematic review and meta-analysis of cognitive behavioural informed psychological interventions for psychiatric inpatients with psychosis. Schizophrenia research, 222, 133–144.

Wood, L., Jacobsen, P., Ovin, F., & Morrison, A. P. (2022a). Key components for the delivery of cognitive behavioral therapies for psychosis in acute psychiatric inpatient settings: a Delphi study of therapists’ views. Schizophrenia Bulletin Open, 3(1), sgac005.

Wood, L., Williams, C., Pinfold, V., Nolan, F., Morrison, A. P., Morant, N., Lloyd-Evans, B., Lewis, G., Lay, B., Jones, R., Greenwood, K., & Johnson, S. (2022b). Crisis-focused Cognitive Behavioural Therapy for psychosis (CBTp) in acute mental health inpatient settings (the CRISIS study): protocol for a pilot randomised controlled trial. Pilot and feasibility studies, 8(1), 205. 10.1186/s40814-022-01160-7

Wood, L., Rowe, A., Persaud, K., Nyikavaranda, P., Nira Malde-Shah, Guerin, E., Dare, C., Constant, C., & Birken, M. (2023). Reflections on the coproduction of a crisis-focused intervention for inpatient settings underpinned by a Cognitive Behavioural Therapy for psychosis (CBTp) model. 1–12. 10.1080/17522439.2023.2220373

Wood, L., Morrison, A. P., Lay, B., Williams, C., & Johnson, S. (2024a). Delivering a Cognitive Behaviour Therapy for psychosis (CBTp) informed crisis intervention in acute mental health inpatient settings: a therapy protocol. Psychosis, 1-12.

Wood, L., Butterworth, H., Nyikavaranda, P., Aderayo Ariyo, Nira Malde-Shah, Guerin, E., Birken, M., Persaud, K., Dare, C., Morant, N., & Johnson, S. (2024b). Patient’s Experiences of a Cognitive Behaviour Therapy Informed Crisis Intervention for Psychosis Delivered in Inpatient Settings: A Qualitative Exploration. Clinical Psychology & Psychotherapy, 31(4). 10.1002/cpp.3033

Wood, L., Morrison, A. P., Birken, M., Dare, C., Guerin, E., Nyikavaranda, P., Nira Malde-Shah, Persaud, K., Ford, P., Cyntheia Nebo, Clarke, C. S., Brynmor Lloyd-Evans, Greenwood, K., Lewis, G., Lay, B., MacLennan, G., Morant, N., Nolan, F., Pinfold, V., & Christiansen, C. (2025). Examining the feasibility of a crisis-focused Cognitive Behaviour Therapy (CBT)–informed psychological intervention for inpatients experiencing psychosis (the CRISIS study): results from a pilot randomised controlled trial. EClinicalMedicine, 86, 103380–103380. 10.1016/j.eclinm.2025.103380

